# Advancing Genotype-Phenotype Analysis through 3D Facial Morphometry: Insights from Cri-du-Chat Syndrome

**DOI:** 10.1101/2025.06.01.25327945

**Authors:** Michiel Vanneste, Harold Matthews, Yoeri Sleyp, Peter Hammond, Mark Shriver, Seth M. Weinberg, Mary L. Marazita, Susan Walsh, Benedikt Hallgrímsson, Ophir D Klein, Richard A Spritz, Kris Van Den Bogaert, Peter Claes, Hilde Peeters

**Affiliations:** Department of Human Genetics, KU Leuven, Leuven, Belgium; Medical Imaging Research Centre, University Hospitals Leuven, Leuven, Belgium; Department of Anthropology, Penn State University, University Park, PA, USA; Center for Craniofacial and Dental Genetics, Department of Oral and Craniofacial Sciences, School of Dental Medicine, University of Pittsburgh, Pittsburgh, PA, USA; Department of Human Genetics, School of Public Health, University of Pittsburgh, Pittsburgh, PA, USA; Department of Biology, Indiana University Purdue University Indianapolis, Indianapolis, IN, USA; Department of Cell Biology & Anatomy, Cumming School of Medicine, University of Calgary, Calgary, AB, Canada; Alberta Children’s Hospital Research Institute, Cumming School of Medicine, University of Calgary, Calgary, AB, Canada; McCaig Bone and Joint Institute, Cumming School of Medicine, University of Calgary, Calgary, AB, Canada; Department of Pediatrics, Cedars-Sinai Guerin Children’s, Los Angeles, CA 90048, USA; Department of Pediatrics, University of Colorado School of Medicine, Aurora, CO, USA; Center for Human Genetics, University Hospitals Leuven, Leuven, Belgium; Department of Electrical Engineering, ESAT/PSI, KU Leuven, Leuven, Belgium

## Abstract

**Purpose:** Facial dysmorphism is a feature of many monogenic disorders, and is important in diagnostics, variant interpretation and nosology. Nevertheless, comprehensively assessing the complex facial shape changes associated with specific syndromes remains challenging. Here, we present 3D morphometric approaches to overcome these limitations, utilizing Cri-du-Chat syndrome (CdCS) as a model.

**Methods:** We analyzed 3D facial photos from 24 individuals with CdCS, 4540 unaffected controls and 5 individuals with rare 5p15.33-15.32 deletions, incorporating two methods to account for age– and sex-related facial variation. We quantified phenotypic variation within and between groups and explored genotype-phenotype correlations in CdCS.

**Results:** We identified changes in the characteristic facial features of CdCS with age and found that facial shape in CdCS differed from controls in highly consistent directions, but with varying magnitudes of effect. 5p15.33-15.32 heterozygotes had non-specific dysmorphic features that were objectively different from those in CdCS, delineating multiple critical regions for facial dysmorphism on chromosome 5p.

**Conclusion:** This work explores 3D facial morphometry to complement the standard clinical assessment of facial dysmorphism. It provides insights into the genetic basis of facial shape in CdCS and highlights the potential of 3D morphometric techniques to facilitate clinical diagnostics, variant interpretation, and delineation of syndrome nosology.

## Introduction

Facial dysmorphism is present in many monogenic disorders, and accurate assessment of dysmorphic features contributes to diagnostics, nosology and variant interpretation^1,2^. While standardized terminology facilitates consistent reporting of dysmorphic features by clinicians worldwide^3^, it remains challenging to capture the complex three-dimensional facial phenotype associated with specific disorders using this lexicon^4^. Furthermore, facial shape is influenced by factors such as age, sex, and genetic ancestry, which complicate evaluating facial dysmorphism and assessing phenotypic variation^1,5^. This study applies 3D morphometric techniques to address these limitations, utilizing Cri-du-Chat syndrome (CdCS; OMIM #123450) as a model.

CdCS is a contiguous gene deletion disorder resulting from heterozygous deletions on chromosome 5p, with an estimated incidence of 1 in 15,000 to 1 in 50,000 live births^6,7^. Ninety percent of cases occur de novo, typically due to terminal deletions arising on the paternal allele^6^, while the remaining ten percent result from unbalanced inheritance of parental structural rearrangements, parental mosaicism or the inheritance of a parental deletion^6,8^. There are no recurring breakpoints, and deletion sizes range from 500 kb to 45 Mb. However, most affected individuals carry terminal deletions of 10 Mb or larger involving the 5p15.31 and 5p15.2 regions^6,8–10^. Typical clinical features of CdCS include global developmental delay, delayed speech development, microcephaly, a high-pitched cat-like cry in infancy and facial dysmorphism^6,11^. However, CdCS demonstrates considerable phenotypic variability, which is in part attributed to differences in deletion size and breakpoints^6,12^. Recurring facial features include hypertelorism, epicanthic folds, a prominent nasal bridge, short philtrum, microcephaly and micrognathia; nevertheless, a recognizable facial gestalt is not always present.

Despite the lack of consistent recurring breakpoints, some genotype-phenotype correlations have been established (Figure 1). The degree of intellectual disability (ID) is associated with the size and extent of the deletion, with usually moderate ID in individuals with deletions limited to 5p15.2 and severe ID in individuals with larger deletions or concurrent chromosomal abnormalities^6,8,10,11^. Similarly, 5p15.32 deletions are linked to speech delay^10^ and deletions of 5p15.31 appear to be critical for the cat-like cry^10,13^. For facial dysmorphism however, there is conflicting evidence about which chromosomal regions are critical. Various overlapping and non-overlapping deletions have been reported in individuals with varying dysmorphic facial features, and the specific contributions of the corresponding chromosomal regions to facial dysmorphism in CdCS thus remain unclear^9–12^.

**Figure 1:**
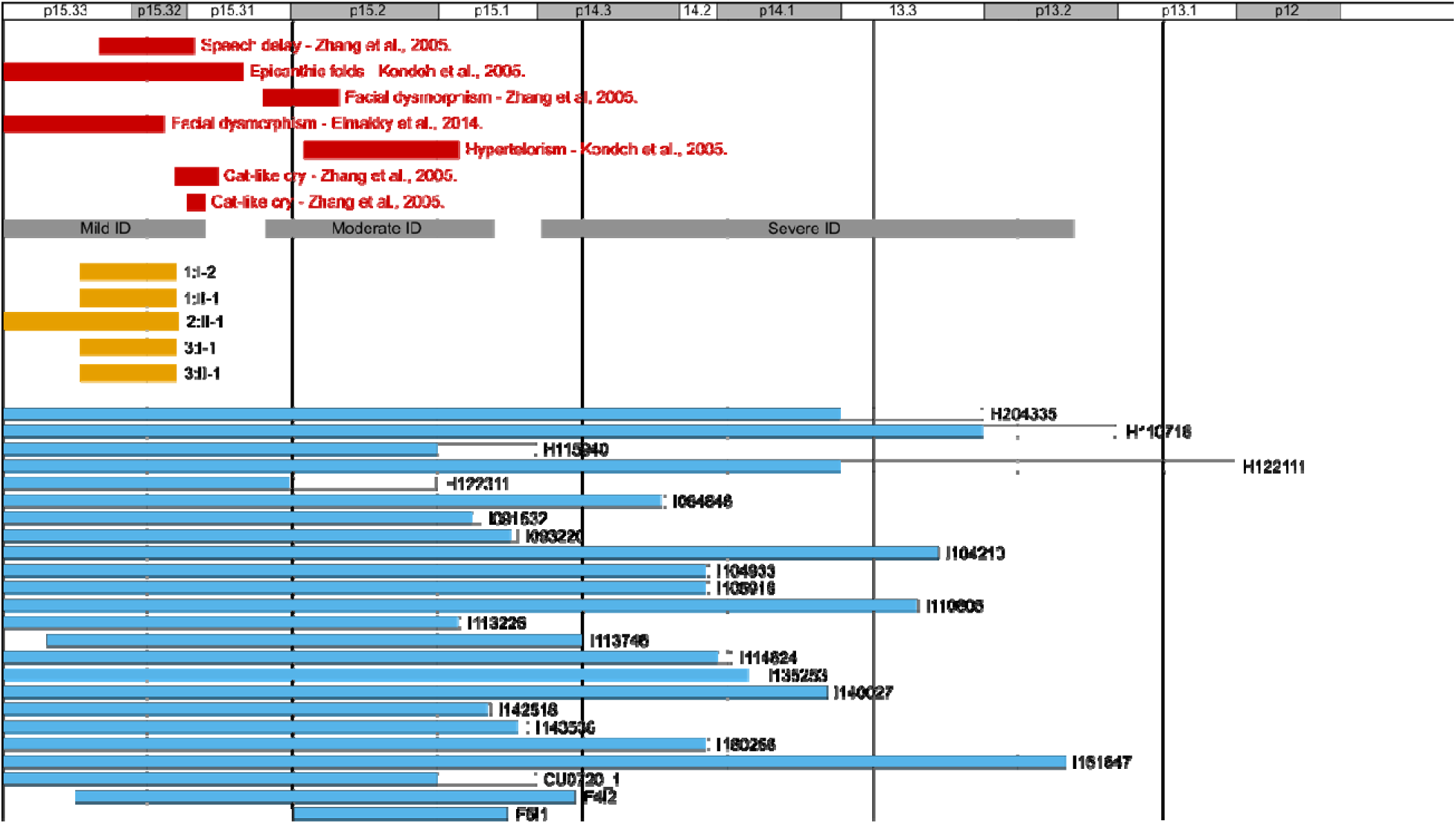
Map of chromosome 5p. Map of chromosome 5p with cytogenetic bands on top, in red the previously reported critical regions for different phenotypic characteristics in Cri-du-Chat Syndrome (CdCS), in grey the genotype-phenotype correlations for intellectual disability and in yellow the deleted regions of the 5p15.33-15.32 deletion heterozygotes. Blue bars represent the minimal size of the deletions in the CdCS reference cohort, the white areas correspond to the maximal size (i.e. last present array probe).

Recently, efforts towards a more objective evaluation of facial dysmorphism have given rise to next-generation phenotyping tools. Advances in morphometric analysis have facilitated the development of tools that can objectively match two– and three-dimensional (3D) photographs to known syndromic gestalts and can suggest syndromic diagnoses^14,15^. The publication of open-source 3D craniofacial growth curves allows clinicians and researchers to evaluate facial morphology independent of age– and sex-related facial features^16^. In addition, recent quantitative approaches to compare facial phenotypes both within and between different molecular diagnoses enable objective delineation of syndrome nosology^1^. Together, these advances make a quantitative, objective assessment of facial dysmorphism possible.

In this work, we used next-generation 3D phenotyping tools to study facial dysmorphism in CdCS. We quantified the phenotypic variation in CdCS, delineated the recurring facial features and investigated the changes in facial phenotype with age. In addition, we also studied the facial dysmorphism in five individuals with small 5p15.33-15.32 deletions and objectively compared their facial phenotype to that of participants with more typical, terminal 5p deletions larger than 10 Mb. By doing so, we delineated critical regions for typical and atypical facial dysmorphism in CdCS.

## Materials and Methods

### Sample composition

We compiled a CdCS reference cohort from 3D facial photos, demographic data (self-reported age, sex, and ancestry), and molecular testing results of 98 participants with a clinical and/or molecular diagnosis of CdCS from three sources. We obtained previously collected data on 59 participants with CdCS from the online FaceBase repository (FB00000861 [https://doi.org/10.25550/TJ0]) and 37 from Peter Hammond’s legacy 3D dysmorphology dataset hosted at KU Leuven. Two participants previously diagnosed with CdCS at the Centre for Human Genetics of University Hospitals Leuven were recontacted for inclusion in this study. We only included participants with available molecular testing results (n=34) and we excluded participants with poor image quality (n=3), incomplete metadata (n=1), additional pathogenic genetic variants besides a 5p deletion (n=1) and self-reported non-European ancestry (n=4) to retain a curated sample of 24 participants with CdCS (Supplementary Data 1). This last exclusion criterion was applied because we corrected for age– and sex-related facial variation using an open-source normative reference for 3D facial shape, which consists solely of participants of recent European ancestry (see *’Growth curves and facial signatures’*)^16^. The genotypic data of the CdCS cohort were determined by different cytogenetic techniques (conventional karyotyping, bacterial artificial chromosome (BAC) comparative genomic hybridization (CGH) array (Agilent) and 180k Cytosure chromosomal microarray analysis (CMA) (Oxford Gene Technologies)), each offering different levels of resolution. All genotype data, including regions of uncertainty due to resolution differences, were mapped to GRCh38 genomic locations using LiftOver from the UCSC Genome Browser (https://genome.ucsc.edu/cgi-bin/hgLiftOver)^17^ and Ensembl (Release 109)^18^ (Figure 1, Supplementary Data 1). The used IDs are not known to anyone outside the research group.

We also collected 3D facial photos and clinical data of five participants with heterozygous 5p15.33-15.32 deletions from three families. All deletions were previously identified through CMA in a clinical work-up at the University Hospital Gasthuisberg (Leuven, Belgium).

We used a large control sample (n=4680) consisting of previously collected 3D facial images and demographic data (age, sex, self-reported ancestry) sourced from the 3D Facial Norms cohort^19^ and studies at the Pennsylvania State University and Indiana University-Purdue University Indianapolis. We excluded participants with ages outside the age range of the 3D craniofacial growth curves (see below), retaining 4540 participants total. Sex and age distribution of this sample is provided in Supplementary Figure 1 and Supplementary Data 2; sample characteristics and participant recruitment have been described in detail by White et al.^20^.

### 3D image acquisition and processing

3D facial images were acquired using two digital stereophotogrammetry systems (3dMDface, Vectra H1). We non-rigidly registered an average facial template to each 3D image using the MeshMonk toolbox (v.0.0.6)^21^ to obtain a standardized facial representation defined by 7160 homologous quasi-landmarks. The deformed template corresponds anatomically across all images and facilitates meaningful comparison between images. We inspected images visually and excluded them if the registration process had failed.

### Growth curves and facial signatures

We calculated facial signatures for all participants, which measure how individual facial shapes deviate from age-and sex-matched normative reference faces^22^. Per participant, we compared all 7160 quasi-landmarks to those of an age– and sex-matched normative reference, determined using open-source 3D craniofacial growth curves^16^. These growth curves provide statistical facial shape models for individuals of recent European ancestry from ages 0.5-68.9 years (females) and 0.5-51.8 years (males); participants not meeting these criteria were necessarily excluded during data curation (see *Sample Composition*). We calculated facial signatures as z-scores for local shape deviation along three Cartesian axes (x-, y-, and z-signatures) as well as locally perpendicular to the facial surface (normal signature), which we used for visualization. Since the control sample was previously included to construct the facial growth curves, we calculated the facial signatures for this sample using a modified approach, comparing each participant’s facial shape to a variation model constructed on the entire sample, excluding their own facial shape. To distinguish between facial shape differences affecting the full face or parts of the face, we performed the assessments described below twice; once including all quasi-landmarks (full face), and once including only the quasi-landmarks that made up the midface (Supplementary Figure 2).

### Quantifying phenotypic variation through facial signatures

We assessed phenotypic variation within and between groups using the methodology described by Matthews et al.^1^. First, we concatenated facial signatures in the x-, y– and z-directions, followed by dimensionality reduction using single-value decomposition. This way each participant’s face is represented by a single feature vector in a high-dimensional ‘feature vector space’ and facial signature similarity is reflected by proximity in that space^23^. For each diagnostic group (CdCS cohort, 5p15.33-15.32del and controls), we calculated a mean feature vector, representing the average facial signature for that group.

The phenotypic similarity between two (individual or mean) facial signatures was then calculated as the cosine similarity between their corresponding feature vectors (cosine similarity = cos(α), with α being the angle between the feature vectors; Figure 2). The cosine similarity ranges from –1 to 1, indicating two feature vectors have exactly opposite or identical directions respectively (thus representing a multivariate correlation between two facial signatures). To measure the severity of a particular phenotype in an individual participant, we projected their individual feature vector onto the feature vector corresponding to that phenotype. The magnitude of the projected vector, also called the ‘severity score’, corresponds to the participant’s phenotypic severity along the direction of the investigated feature vector (Figure 2). We performed pairwise comparisons of the cosine similarities to the CdCS mean and CdCS severity scores between all three groups using a two-sample Kolmogorov-Smirnov test (kstest2, Matlab R2024b).

**Figure 2:**
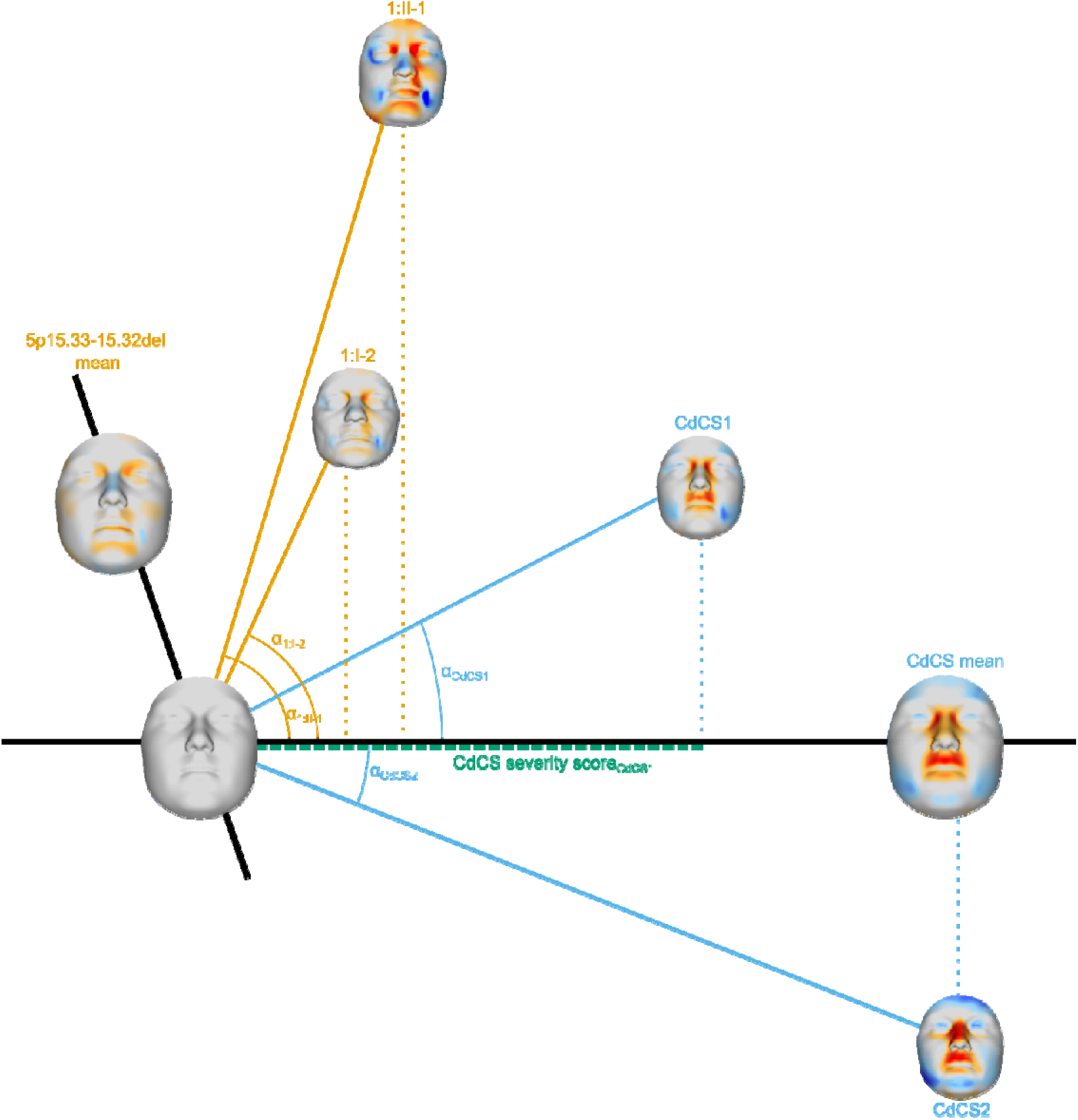
Quantifying phenotypic variation. Two-dimensional representation of the multidimensional ‘face space’ used to quantify phenotypic variation. The mean facial signature of the Cri-du-Chat Syndrome (CdCS) cohort and 5p15.33-15.32del heterozygotes are shown, as well as their corresponding feature vector (black bold lines). The facial shapes and signatures of different participants belonging to both groups (orange = 5p15.33-15.32del; blue = CdCS) are displayed. The cosine similarity to the CdCS mean is calculated based on the angle between the individual and mean CdCS feature vectors (arches), the CdCS severity score is calculated by projecting the individual on the mean feature vectors (dotted lines), as illustrated for CdCS-1 in green.

We quantified within-group phenotypic variation using two metrics: ‘directional variation’ and ‘severity variation’. Directional variation measures the variation in the directions of individual feature vectors within a group, and was computed as the root mean square of 1-cosine similarity between each member and the group mean. Severity variation measures the variation in the magnitudes of individual feature vectors along the direction of the group mean and was computed as the standard deviation of the severity scores. Throughout these calculations, we used a leave-one-out approach where the participant being scored is excluded from the mean signature estimation. To interpret the magnitude of the directional and severity variation values, we compared the results to previously published values from 39 monogenic disorders with facial dysmorphism^1^.

### Statistical shape analysis through covariate-adjusted PCA

We performed principal component analysis (PCA) to assess phenotypic similarity independent from the cosine similarity analysis. We aligned the facial shapes of the control sample using generalized Procrustes analysis and adjusted for sex, age and age-squared using partial least squares regression (PLSR). We then Procrustes superimposed the CdCS cohort shapes onto the mean control shape and corrected them for age, sex and age-squared using the PLSR coefficients from the control sample model to account for general facial variation introduced by these covariates. The aligned and covariate-adjusted shapes were then subjected to PCA in three different designs. We performed PCAs on the CdCS cohort (CdCS-PCA) and control sample (control-PCA) separately to assess within-group variation, and we assessed between-group variation through a joint PCA of the CdCS cohort and a matched control sample of equal size (matched-PCA, Supplementary Data 3). We constructed the matched control sample by, in a random order, selecting individual participants from the larger control sample with matching age and sex to the participants of the CdCS cohort. Once a participant was selected as matched control, they were removed from the pool of possible control picks for the other participants, ensuring the matched control sample consisted of 24 unique individual participants. Lastly, we projected the facial shapes of the 5p15.33-15.32del heterozygotes into the matched-PCA space to evaluate similarity-based clustering for these participants. For all PCAs we used a cutoff of 98% cumulative variance explained.

### Effect of age, sex and deletion size

To investigate whether facial shape was associated with variation in age and deletion size in the CdCS cohort, we performed linear regression (fitlm, Matlab R2024b) between age and deletion size as predictor variables and the CdCS severity scores, cosine similarities to the CdCS mean and principal component (PC) scores of the covariate-adjusted CdCS-PCA as response variables. We adjusted P-values for multiple testing using the Bonferroni correction. To test the robustness of the covariate adjustment in the CdCS cohort, we applied the same adjustments to matched control sets and assessed residual age correlations. In random order, we selected controls with the same sex and the closest matching age to the individual participants of the CdCS cohort to construct matched control test samples (n = 24). We aligned these samples’ facial shapes and adjusted them for sex, age, and age-squared using the coefficients of a PLSR model constructed on the remaining controls (n = 4516), before subjecting the adjusted shapes to PCA. We repeated this process 100 times, each time selecting different controls for the test set, and found no significant associations between age and the PC scores of the covariate-adjusted test set (Supplementary Figure 3, Supplementary Data 4).

## Results

### Cohort description

We assembled a CdCS reference cohort of 24 participants with a heterozygous 5p deletion (Figure 1). Twenty-one deletions were terminal and three were interstitial, and deletion sizes ranged from 7-42 Mb. Importantly, all participants exhibited deletions encompassing the 5p15.2 region, which was previously reported as critical for the ID and facial dysmorphism of CdCS (Figure 1)^10,12^. Detailed molecular characteristics of this cohort are reported in Supplementary Data 1. In addition to this reference cohort, we included five participants from three families with smaller heterozygous 5p15.33-15.32 deletions of 3.3 Mb and 6.0 Mb (Figure 1, Figure 3). Molecular karyotyping in these families was initially performed because of facial dysmorphism and delayed motor development in participant 1:II-1 and because of developmental delay in participants 2:II-1 and 3:II-1. Table 1 contains the primary clinical and molecular data of these participants; detailed clinical data are available upon request to the authors.

**Figure 3:** 5p15.33-15.32 deletion heterozygotes. **(a)** Pedigrees of the 5p15.33-15.32 deletion heterozygotes. **(b)** Facial photographs of the five 5p15.33-15.32 deletion heterozygotes. All participants gave permission for publication of these pictures. **(c)** Facial signatures of the five 5p15.33-15.32 deletion heterozygotes. The colors correspond to the degree of local shape deviation (in Z-scores compared to an age– and sex-matched normative reference) locally perpendicular to the surface (blue = inward displacement, red = outward displacement). For participant 3:I-1 no facial signature was calculated, as their images did not pass quality control due to their beard. **All panels of this figure contain potentially identifiable characteristics and have been removed in this preprint. Access to these materials can be requested by contacting the corresponding author.**

**Table 1:** Summary of primary data for 5 participants with 5p15.33-15.32 deletion.

| ID | Age at inclusion | Sex | High pitched voice as infant | Developmental delay (DD) | Micro-cephaly | Speech delay | Molecular (breakpoints) results and inheritance |
| --- | --- | --- | --- | --- | --- | --- | --- |
| 1:I-2 | 46-50 | Female | Unknown | No | No | No | 46, XX, del 5p15.33-5p15.32 (2682974-5982763), inheritance unknown |
| 1:II-1 | 10-15 | Male | Yes | Motor delay* | No | No | 46, XY, del 5p15.33-5p15.32 (2682974-5982763), maternally inherited |
| 2:II-1 | 16-20 | Male | Yes | Mild global DD | No | No | 46, XX, del 5p15.33-5p15.32 (16497-6093434), de novo |
| 3:I-1 | 56-50 | Male | Unknown | Mild global DD | No | No | 46, XY, del 5p15.33-5p15.32 (2682974-5982763), inheritance unknown |
| 3:II-1 | 30-35 | Female | No | Yes | No | Yes | 46, XX, del 5p15.33-5p15.32 (2682974-5982763), paternally inherited |

### Facial signatures reliably assess individual and recurring dysmorphic features

Clinical assessment of the CdCS reference cohort showed that all participants had some facial features typical for CdCS, such as short philtrum (24/24; 100%), prominent nasal bridge (21/24; 88%), epicanthic folds (20/24; 83%), micrognathia (17/24; 71%) and hypertelorism (12/24; 50%) (Figure 4A, Supplementary Data 1). In only 7/24 participants (29%) all five typical features were present, while 18/24 participants (75%) had at least four of these features. Clinically, two participants (8%; I161647 and I091532) lacked the characteristic facial gestalt typically associated with CdCS. Both had the characteristic short philtrum and one had epicanthic folds, and both also had coarser facial features but without prominent nasal bridge, micrognathia or hypertelorism (Supplementary Figure 4, Supplementary Data 1).

**Figure 4:**
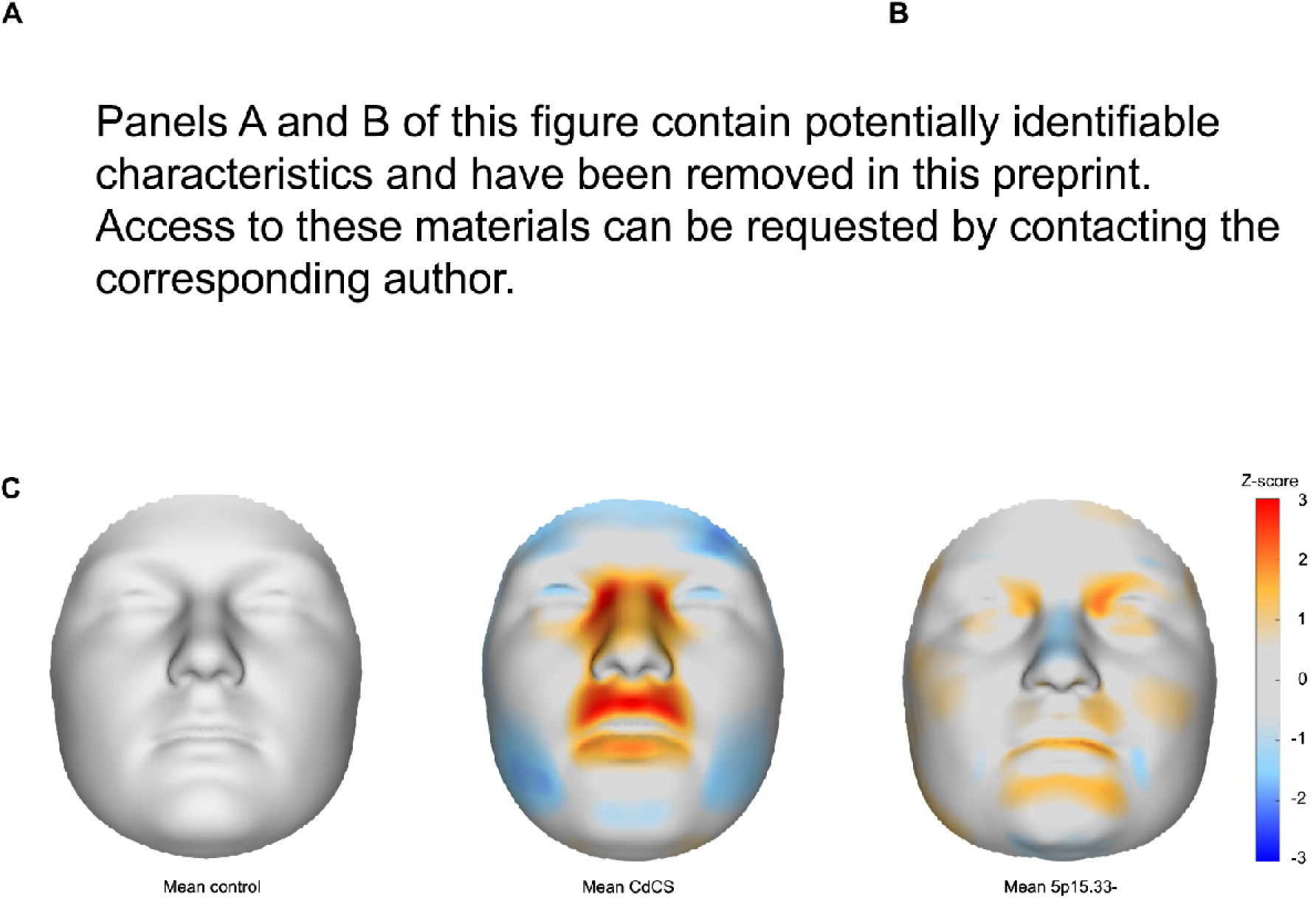
Facial dysmorphism in CdCS. **(a)** Participant from the Cri-du-Chat Syndrome (CdCS) cohort with typical facial features of CdCS, including a prominent nasal bridge, short philtrum, hypertelorism and micrognathia. Written informed consent for recognizable publication was provided. **(b)** Facial signature of the same participant, highlighting the typical dysmorphic features of CdCS. Red indicates outward deviation locally perpendicular to the surface, blue indicates inward deviation. **(c)** Mean facial shapes and signatures for the control sample, CdCS reference cohort and 5p15.33-15.32del heterozygotes.

We calculated facial signatures for all participants in the CdCS cohort to perform an objective assessment of facial shape on an individual and group level (Figure 4B). Individually, facial signatures highlight how a participant’s facial shape deviates from an expected normative face for their age and sex. There was high concordance between the facial signatures and the clinical assessment for the nasal bridge and philtrum, and lower concordance for micrognathia. In three participants, micrognathia was clinically subtle but clear in the facial signature, while in another three a clinical impression of micrognathia was not confirmed by the facial signature. Evaluating the concordance between both assessments for epicanthic folds and hypertelorism was not possible, as these features cannot be distinguished from each other on a facial signature. The mean facial signature of the CdCS cohort (Figure 4C) showed a short philtrum, prominent nasal bridge, triangular facial shape with micrognathia, and a strong signal at the inner canthi suggestive of epicanthic folds, hypertelorism or both. The mean facial signature for the control sample (n = 4540) showed no recurring features (Figure 4C). These findings align with the clinical assessment and previous reports^6,11^, demonstrating how mean facial signatures can visualize the recurring facial features in a cohort.

### Cosine similarity provides quantitative assessment of between-group variation

To quantify facial variation between the CdCS cohort and control sample, we used two metrics that separate phenotypic variation into two components: cosine similarity and the severity score. First, we used the facial signatures to construct a multidimensional space, where each individual is represented by a lower-dimensional feature vector and proximity in the space corresponds to similarity in facial signature. The cosine similarity measures the similarity in direction of two feature vectors, and the severity score measures the magnitude of the feature vectors along a specific direction (Figure 2). The participants of the CdCS cohort had significantly higher cosine similarities to the CdCS mean than controls for both the full face and the midface, indicating that the direction of their feature vectors was consistently different from controls (p = 4.0×10^−15^ and p = 1.1×10^−11^, respectively; two-sample Kolmogorov-Smirnov test; Figure 5A-B, Supplementary Data 6). There was no association between a participant’s deletion size or age and their cosine similarity to the CdCS mean or CdCS severity score (Supplementary Figure 5). We performed a second, independent assessment of between-group variation through PCA on the facial shapes of the CdCS cohort and a matched control sample (PCA_matched_). PC1 and PC2 were both strongly correlated with the CdCS cosine similarity and severity scores (Supplementary Figure 6) and captured core clinical characteristics of CdCS such as the short philtrum, prominent nasal bridge, hypertelorism and micrognathia (Figure 5C, Supplementary Media 1 and 2). The PC1 and PC2 scores for participants with CdCS varied considerably but could still be used to distinguish cases from controls (Figure 5C), confirming the findings of the cosine similarity analysis that the facial phenotype in CdCS is variable but consistently different from controls.

**Figure 5:**
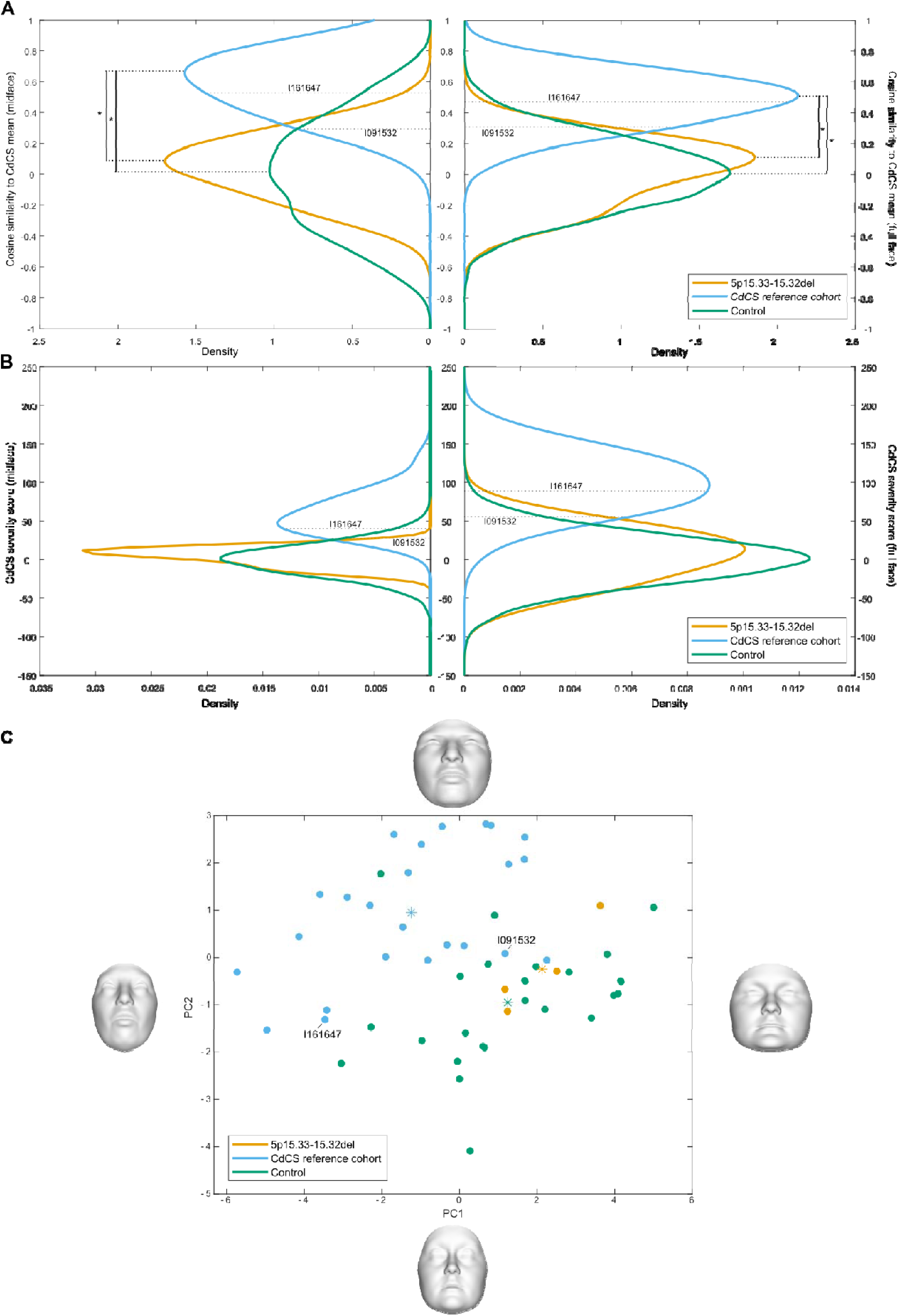
Between-group phenotype comparisons. Probability density functions of the cosine similarity to the Cri-du-Chat Syndrome (CdCS) mean **(a)** and CdCS severity scores **(b)** for the participants of the CdCS reference cohort, controls and the 5p15.33-15.32del heterozygotes, calculated on the full face (right) and midface (left). In **(a)**, significant differences between density functions are marked with an asterisk (Two-sample Kolmogorov-Smirnov test, Bonferroni correction). **(c)** Scatter plot of the first two principal components of the PCA on the CdCS cohort and matched controls. 5p15.33-15.32del heterozygotes were projected into the PC space., stars indicate the projected positions of the group means. The facial shapes show the facial shape at +- 3 standard deviations of the PC scores. In all panels, the scores for the two participants with a clinical outlier phenotype are indicated.

### Facial signatures in CdCS have a consistent direction but variable magnitude of effect

We quantified within-group phenotypic variation by aggregating the cosine similarity and severity scores into respective ‘directional variation’ and ‘severity variation’ scores per group, calculated separately for the full face and the midface. Directional variation quantifies differences in the pattern of how facial features covary, while severity variation describes differences in the magnitude of facial features. Full face directional variation in the CdCS cohort was in the 9th lowest percentile (0.54; 95% confidence interval (CI) = [0.45-0.58]) compared to other monogenic disorders^1^. The midface, which includes most of the characteristic dysmorphic features of CdCS, showed even lower directional variation (0.45; CI = [0.35, 0.45]), whereas directional variation was significantly higher in controls (full face = 1.01; CI = [1.00, 1.02], midface = 1.04; CI = [1.02,1.05]). This means the pattern of covariation of phenotypic features in CdCS is very consistent, especially in the midface. In contrast, the full-face severity variation was low for both CdCS and controls (CdCS severity variation = 35.4; CI = [27.5, 49.7], control severity variation = 30.4; CI = [29.7, 31.0]), scoring in the 7th and 3rd lowest percentiles, respectively. Interestingly, the CdCS midface severity variation was significantly higher than in controls (CdCS severity variation = 26.7; CI = [20.8, 37.5], control severity variation = 17.2; CI = [16.8, 17.5]), indicating the facial signatures in the midface are highly consistent in direction but variable in magnitude.

### The typical facial features of CdCS change with age

We performed a PCA on the CdCS facial shapes (CdCS-PCA) to investigate the primary axes of phenotypic variation within this cohort. PC1 was significantly associated with age even though the facial shapes were adjusted for general facial variation associated with age, corroborating previous reports that the facial features of CdCS change as individuals get older^8,24^ (Supplementary Figure 7A). The shape effects captured by PC1 (Supplementary Figure 7A) showed a more prominent nasal bridge and micrognathia as individuals get older, and less pronounced hypertelorism and/or epicanthic folds. The philtrum, which was consistently assessed as short in all participants in the CdCS cohort, showed little to no change with increasing age. The same effects were present when comparing the mean facial signatures of the youngest twelve CdCS participants (ages 2-12 years; mean 6.5 and median 6.4 years) to that of the oldest twelve participants (ages 15-43 years, mean 22 and median 19 years) (Supplementary Figure 7C-D). A covariate-adjusted PCA of facial shape in the control sample (control-PCA) did not show significant associations of any PC with age and showed different primary axes of variation (Supplementary Figure 7B), indicating that the age-associated axis of variation is specific to the CdCS cohort. Linear regression of the covariate-adjusted CdCS shapes on age also indicated a changing phenotype with age (Supplementary Media 3); however, broad generalizations are challenging due to the uneven age distribution in our cohort.

### Objective facial analysis explains outlier phenotypes

Two participants of the CdCS cohort did not exhibit the typical CdCS facial gestalt on clinical assessment (I091352 and I161647), and we attempted to quantify what facial features define the outlier clinical impressions. For both participants, the facial signatures corresponded well to the clinically described coarser facial features and the absence of a prominent nasal bridge and epicanthic folds (Supplementary Figure 4A). For I091352, objective analysis confirmed the clinical assessment that their facial shape was atypical for CdCS. They had a low cosine similarity to the CdCS mean, a low CdCS severity score and clustered closely to controls on the matched-PCA (Figure 5A-C). For I161647, however, the cosine similarity to the CdCS mean and CdCS severity score were close to the average of the CdCS cohort, suggesting a relatively typical CdCS phenotype (Figure 5A-B). This individual was the oldest participant of the CdCS cohort. While their phenotypic features were not atypical for CdCS, they were atypical for someone with CdCS at their age (Supplementary Figure 8). The clinical impression of an outlier phenotype in this participant was thus likely explained by their less prominent nasal bridge and absence of micrognathia on a background of normal age-related variation, which is an unusual combination among older individuals with CdCS.

### Facial features in 5p15.33-15.32del heterozygotes are different from those in typical CdCS

Finally, we compared the facial features of the 5p15.33-15.32del heterozygotes to the CdCS reference cohort, who have ‘typical’ deletions involving 5p15.2, to assess the contribution of these distinct 5p regions to the facial dysmorphism in CdCS. Clinically, only participant 1:II-1 had some facial features that were reminiscent of typical CdCS, with hypertelorism and epicanthic folds (Figure 2B). The other 5p15.33-15.32del heterozygotes showed no marked facial dysmorphism and no recurring dysmorphic features. Their facial signatures (Figure 2C) highlighted the facial asymmetry and hypertelorism in participant 1:II-1 and mild hypertelorism in participants 1:I-2 and 2:II-1, and no participant had the prominent nasal bridge and short philtrum of the CdCS mean signature. The mean signature of the 5p15.33-15.32del heterozygotes (Figure 3C) suggested these individuals have a similarly shaped nasion and a prominent chin; however, this impression must be interpreted cautiously because of the small sample size. The 5p15.33-15.32del heterozygotes had significantly lower cosine similarities to the CdCS mean than the CdCS reference cohort (p = 0.0010, two-sample Kolmogorov-Smirnov test), and their scores did not differ significantly from those of controls (p=0.93) (Figure 5A). When projected into the matched-PCA space, the 5p15.33-15.32del heterozygotes clustered with controls rather than with the CdCS cohort, again indicating their facial features are different from individuals with typical 5p deletions.

## Discussion

Accurately assessing dysmorphic facial features can contribute to correct diagnosis, accurate nosology and variant interpretation in clinical genetics, but this can be challenging even for experienced dysmorphologists^5,25^. In this work, we apply 3D morphometric techniques to objectively study facial dysmorphism in CdCS. We calculated facial signatures to account for age– and sex-related variation and found they were highly concordant with the clinical assessment on an individual level. In some younger participants with CdCS, a finding of micrognathia was not present on the facial signature. This discrepancy could be attributable to prior knowledge about the CdCS diagnosis influencing the assessment (i.e., confirmation bias), as clinical re-evaluation of these participants showed they indeed did not exhibit micrognathia. On a group level, the mean facial signature showed that the most consistent facial features of CdCS are hypertelorism, prominent nasal bridge, short philtrum, and prominent upper lip. These findings are in line with the clinical literature^6,9,12^, and we consider the mean facial signature as a valid representation of the CdCS facial gestalt. In addition to previous reports^1^, this work illustrates that individual and mean facial signatures can objectively capture complex syndromic shape transformations and can assist clinicians in interpreting facial dysmorphism.

Quantitative assessment of phenotypic variation revealed a highly consistent direction of facial shape change in the participants with typical 5p deletions. This may seem to contrast with the reported phenotypic variability in CdCS^6,11^, but in reality highlights the complementarity of accurate quantitative 3D phenotyping and traditional clinical assessments. While the direction of facial shape change associated with CdCS was highly consistent, the magnitude of this shape change was variable, especially in the midfacial region. The direction of effect and effect size of a facial shape transformation are likely difficult to discern clinically, and this may contribute to the clinical perception of phenotypic variability in CdCS. Furthermore, variation in age and sex also contribute to phenotypic variability and complicate the clinical assessment, while the approach presented here corrects for these sources of variation using craniofacial growth curves^16^. Lastly, the presence or absence of minor qualitative features like epicanthic folds can influence a dysmorphologist’s opinion in comparing patients^25^. In facial signatures, however, minor qualitative features that involve few data points have less influence on the global shape transformation than shape changes that involve many data points, such as a prominent nasal bridge and short philtrum in this example. The 3D morphometric tools presented here thus allow for a comprehensive quantitative assessment of phenotypic variability that complements clinical assessment.

We used the morphometric tools to complement the clinical assessment of a male patient (participant 1:II-1) with motor delay, pronounced hypertelorism and epicanthic folds. Genetic testing revealed a small heterozygous 5p15.33-15.32 deletion, and the dysmorphic features were clinically interpreted as consistent with the typical CdCS facial phenotype. However, their carrier parent had no dysmorphic facial features. Morphometric analyses in a cohort of five 5p15.33-15.32 deletion heterozygotes showed no consistent dysmorphic features, either on clinical assessment or through facial signatures. Their facial signatures were different from typical CdCS, and their cosine similarity to CdCS was as low as that of controls. These findings illustrate one of the pitfalls of clinically assessing facial dysmorphism, where certain features, such as epicanthic folds in this case, can be overinterpreted despite the rest of the facial phenotype matching poorly. This overinterpretation is especially likely when facial features are reinterpreted after molecular results are known, which will likely become more common with the increase in genotype-first approaches^26^. In these cases, 3D facial phenotyping can assist clinicians and molecular geneticists in remaining objective and resolving diagnostic questions, as an added computer-based dimension to the clinical art of dysmorphology.

Previous studies have been inconclusive about which chromosomal regions are critical for facial dysmorphism in CdCS^10,12^. Here, we found compelling evidence for the existence of multiple critical regions for facial dysmorphism on chromosome 5p. Individuals with deletions limited to 5p15.33-15.32 presented with heterogeneous facial dysmorphism that was different from the typical CdCS features in individuals with deletions including 5p15.2. This suggests the existence of a 3 Mb region on 5p15.33-15.32 for which deletion causes atypical facial dysmorphism that can include epicanthic folds, and a second region on 5p15.2 that is critical for features such as ocular hypertelorism, prominent nasal bridge and short philtrum. This could explain the conflicting previously reported regions, as well as the occurrence of atypical phenotypes in individuals with small terminal or subterminal deletions^9,10,12,27^. We found no other correlations between deletion size and phenotypic severity or typicality, suggesting that regions centromeric to 5p15.2 do not contribute to the facial dysmorphism of CdCS and that other factors such as DNA methylation and polygenic background also contribute to facial phenotypic variation in CdCS^28,29^.

This work illustrates the strength of 3D morphometry in clinical genetics research. Compared to 2D photography, 3D imaging captures more anatomical data. This enables detailed analyses even in studies with limited sample sizes, an important advantage when investigating rare diseases. However as with many emerging technologies, 3D morphometry still faces methodological challenges. While craniofacial growth curves have improved the robustness and usability of 3D phenotyping^16,30^, they are currently limited to individuals of European ancestry. Second, the scarcity of 3D images of individuals with rare conditions increases the risk of ascertainment bias^31,32^. Expanding access to 3D facial imaging across diverse populations could overcome these limitations^33^. Lastly, the current dense surface registration toolbox limits the quantitative analysis strictly to facial shape, meaning other dysmorphic features that are frequent in CdCS such as microcephaly and preauricular tags could not be assessed^6,11^. Despite these limitations, our findings demonstrate that 3D morphometrics can be used to answer important clinical and research questions. We applied these tools to assess genotype-phenotype correlations, a key challenge in clinical genetics. 3D morphometrics can also assist in diagnostics and variant interpretation^34^, as well as provide insights into the genetics of facial shape^20,29,35^. In conclusion, 3D facial morphometry offers an objective, quantitative assessment of facial features that is complementary to the standard qualitative clinical assessment.

## Data Availability

Phenotype, genotype and demographic data of the CdCS cohort were mostly collected previously and were obtained from the online FaceBase repository (FB00000861 [https://doi.org/10.25550/TJ0]) and an in-house collection. Access to the FaceBase data requires institutional ethics approval and approval from the FaceBase data access committee. Access to the 3D facial surface models of the previously existing in-house collection of participants with CdCS (n= 37) is not possible because these data were previously collected without consent for broad data sharing consent. Access to the facial surface models and clinical data that were collected for this study (two participants with CdCS, five participants with a 5p15.33-15.32del and their relatives) is not possible because consent for data sharing was not provided.

The control sample (n = 4680) was collected previously and included samples from the 3D Facial Norms cohort and studies at the Pennsylvania State University and Indiana University-Purdue University Indianapolis. The 3D facial surface models of the 3D Facial Norms cohort are available through the FaceBase Consortium (FB00000491.01 [https://doi.org/10.25550/VWP]). The participants making up the PSU and IUPUI datasets were previously collected without consent for broad data sharing. This restriction is not because of any personal or commercial interests. Additional details can be requested from M.D.S. and S.W. for the PSU and IUPUI datasets, respectively.

The MeshMonk toolbox for mesh-to-mesh registration is publicly available at https://gitlab.kuleuven.be/mirc/meshmonk.

## Author Contributions

Conceptualization: M.V., H.P.; Data curation: M.V., H.M.; Software: M.V., H.M., P.C.; Formal analysis: M.V., K.V., H.P.; Visualization: M.V.; Funding acquisition and resources: P.H., M.S., S.M.W., M.L.M., S.W., S.R., B.H., O.D.K., R.A.S., P.C., H.P.; Supervision: P.C., H.P.; Writing-original draft: M.V., H.P.; Writing-review & editing: M.V., H.M., Y.S., P.H., M.S., S.M.W., M.L.M., S.W., S.R., B.H., O.D.K., R.A.S., K.V., P.C., H.P.

## Ethics Declaration

This study was approved by the ethical review board of KU Leuven and University Hospital Leuven (S65629, Leuven, Belgium). All participants or their legal representatives gave written informed consent prior to participation. Informed consent for publication of recognizable images was obtained and archived from all participants whose recognizable images are included in this study. Local institutional approval was granted for access to the FaceBase Repository (S60658, Leuven, Belgium). The previously collected human data used in this study was collected in different centers under the appropriate local ethical approvals, and all participants gave written informed consent prior to participation.

## Supporting information

Supplementary Media 1-3

## Supplements

**Supplementary Figure 1:**
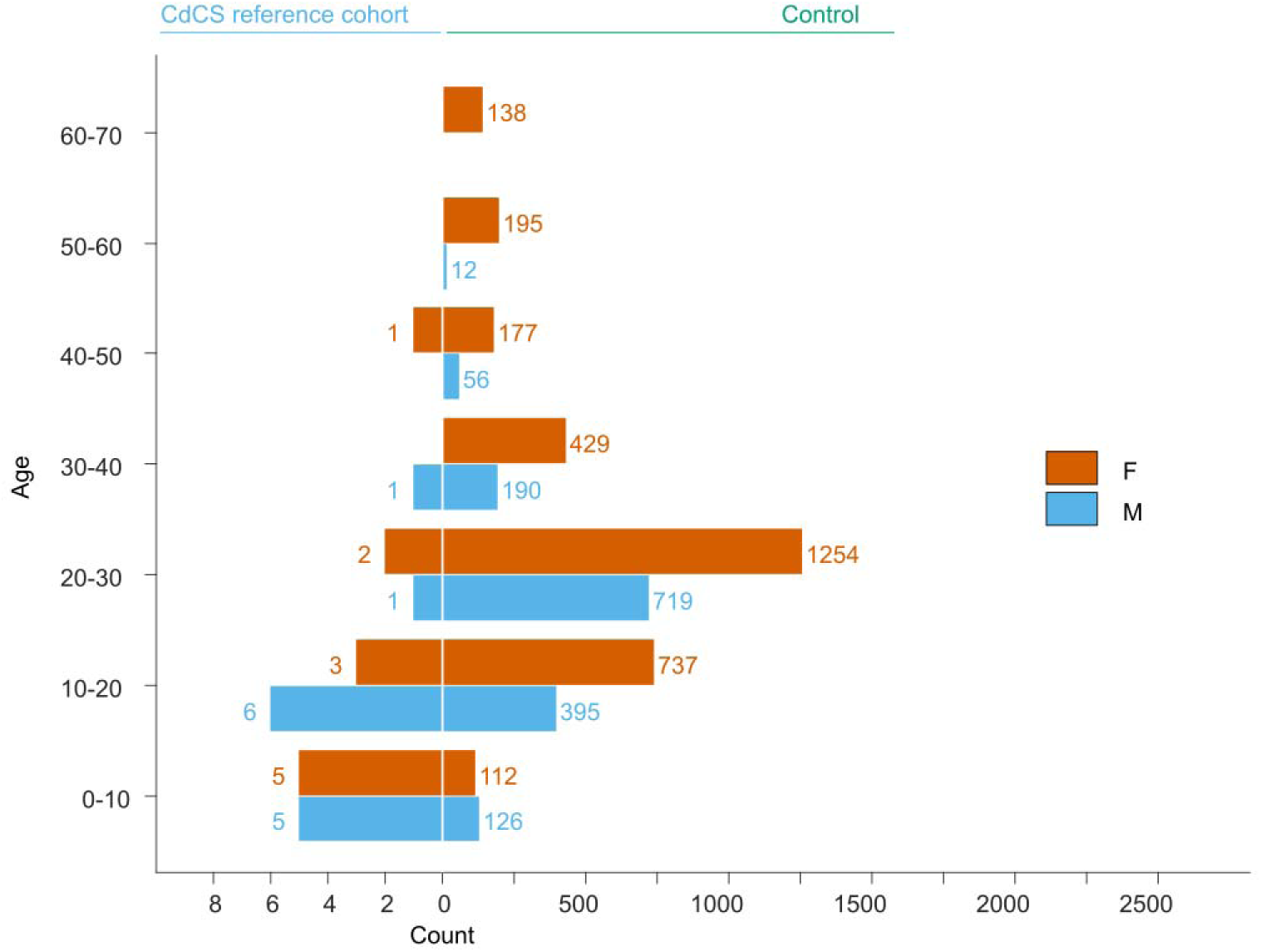
Sample characteristics. Age and sex distribution of the Cri-du-Chat Syndrome (CdCS) cohort and control sample.

**Supplementary Figure 2:**
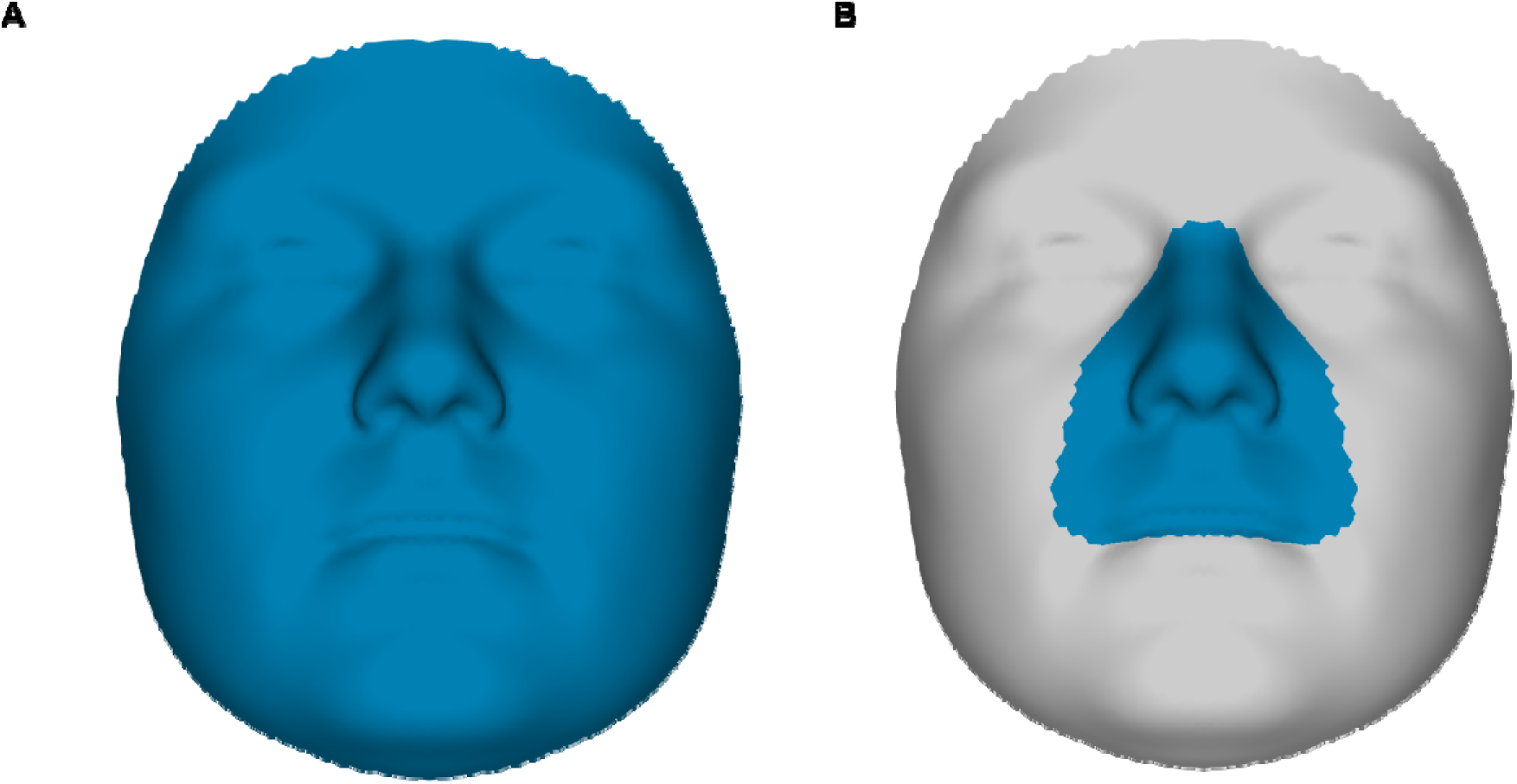
Facial segments. Morphs showing in blue the parts of the face that are included in the analysis of the **(a)** full face and **(b)** midface.

**Supplementary Figure 3:**
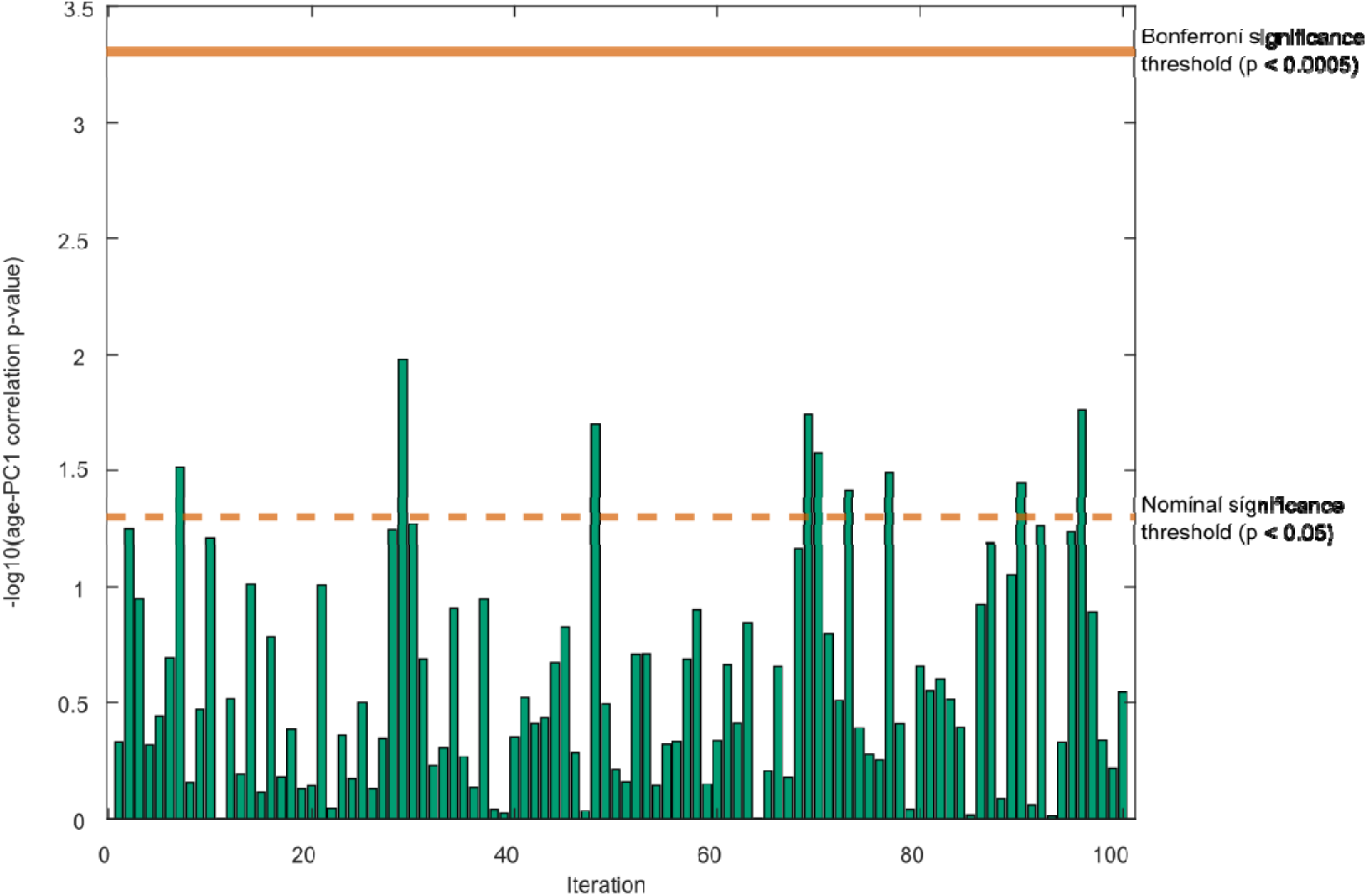
Age correction. Residual correlation between age and the covariate-adjusted shapes of a matched control test sample (n = 24), to test of the robustness of the covariate adjustment of the CdCS cohort. Bars represent the two-tailed –log10 p-values of linear regressions between age and the first principal component (PC1) of the covariate-adjusted facial shapes of the test sample, for 100 iterations with each time different matched controls. Data on other PCs is provided in Supplementary Data 4. The bold line indicates the Bonferroni-corrected significance threshold, the dotted line the nominal significance threshold.

**Supplementary Figure 4:**
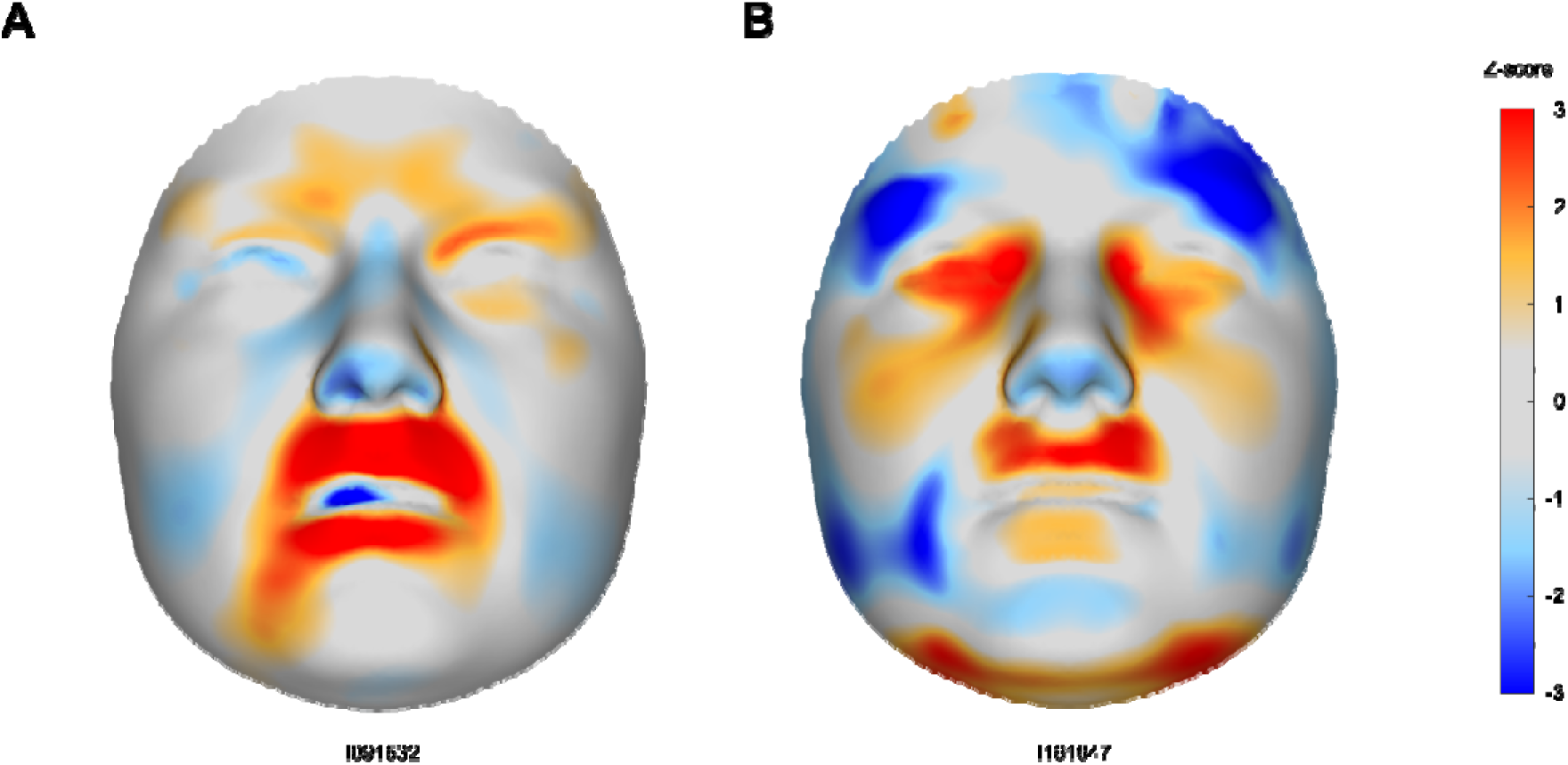
Atypical facial gestalt. Facial signatures of the two participants of the Cri-du-Chat Syndrome (CdCS) cohort with a clinical assessment of an atypical facial gestalt for CdCS. Red indicates outward deviation locally perpendicular to the surface, blue indicates inward deviation.

**Supplementary Figure 5:**
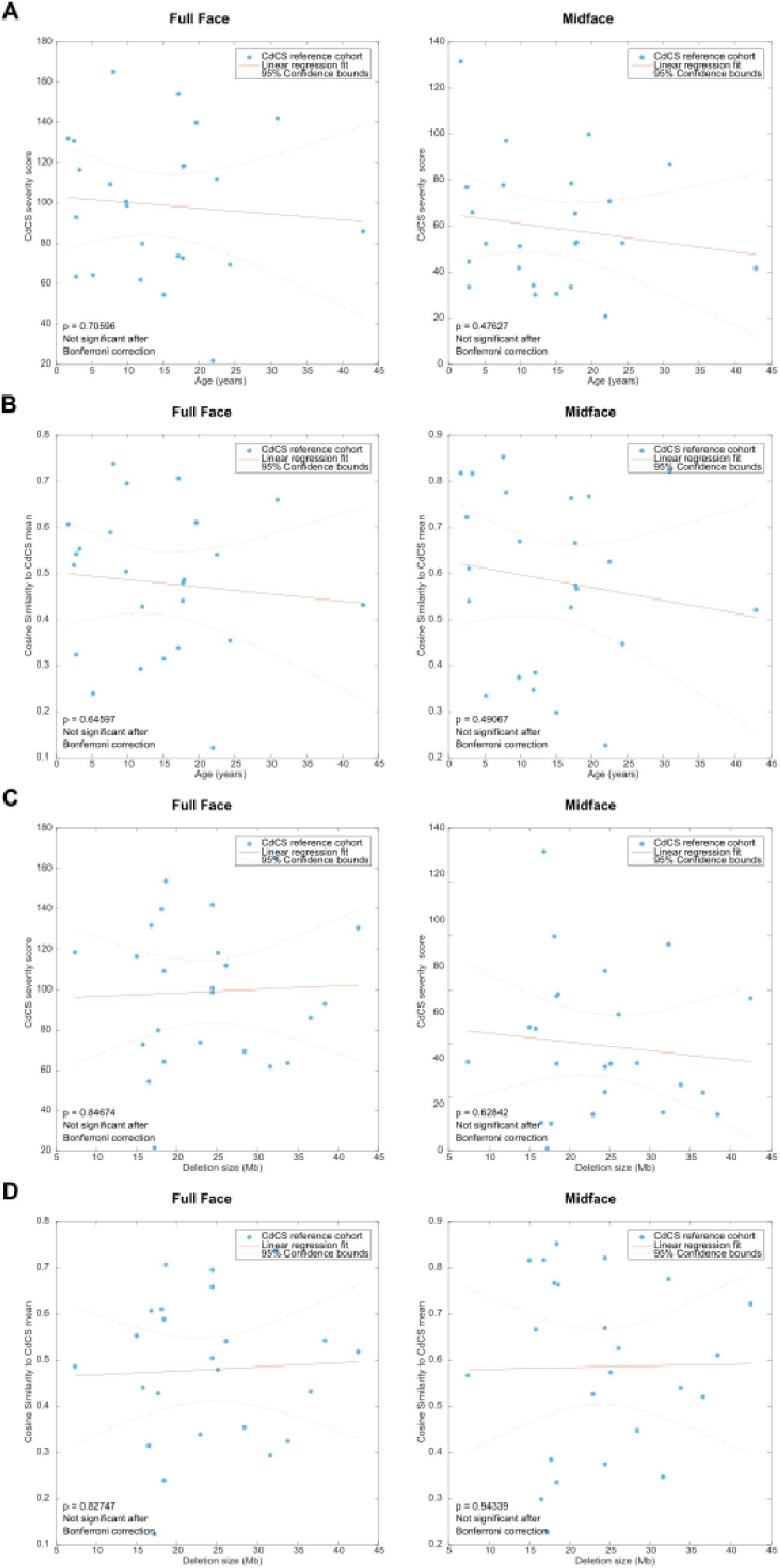
Linear regression of age and deletion size on phenotyping scores. Linear regressions of age and deletion size on the Cri-du-Chat Syndrome (CdCS) severity score **(a, c)** and cosine similarity to the CdCS mean **(b, d)** for the participants of the CdCS cohort. The left panels contain the regressions for the phenotyping scores on the full face, the right panels for the phenotyping scores on the midface.

**Supplementary Figure 6:**
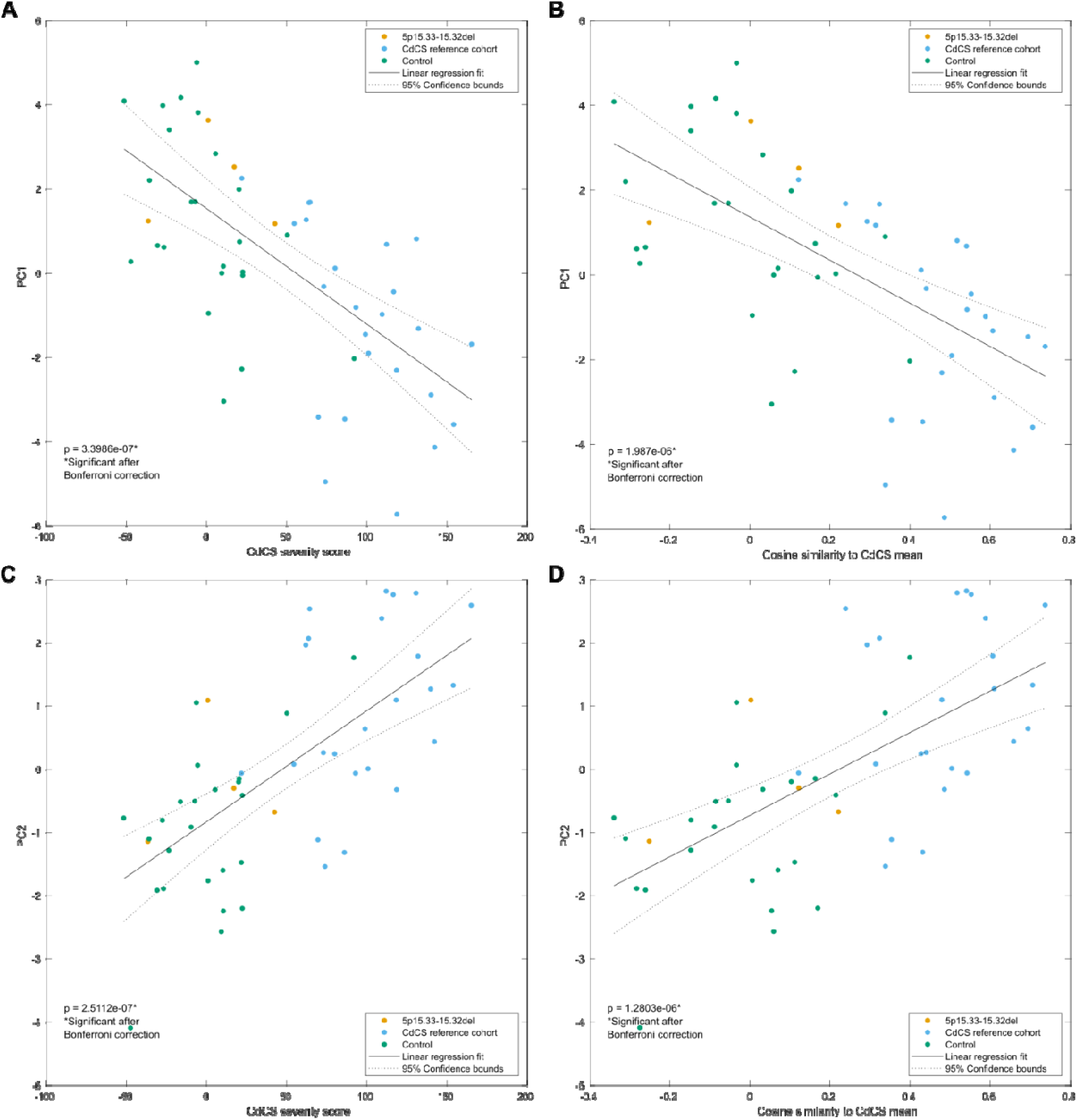
Linear regression of phenotypic scores on PCs. Linear regression of the first two principal components of the PCA on the Cri-du-Chat Syndrome (CdCS) cohort and matched controls versus the CdCS severity score **(a, c)** and cosine similarity to the CdCS mean **(b, d)**, calculated on the full face.

**Supplementary Figure 7:**
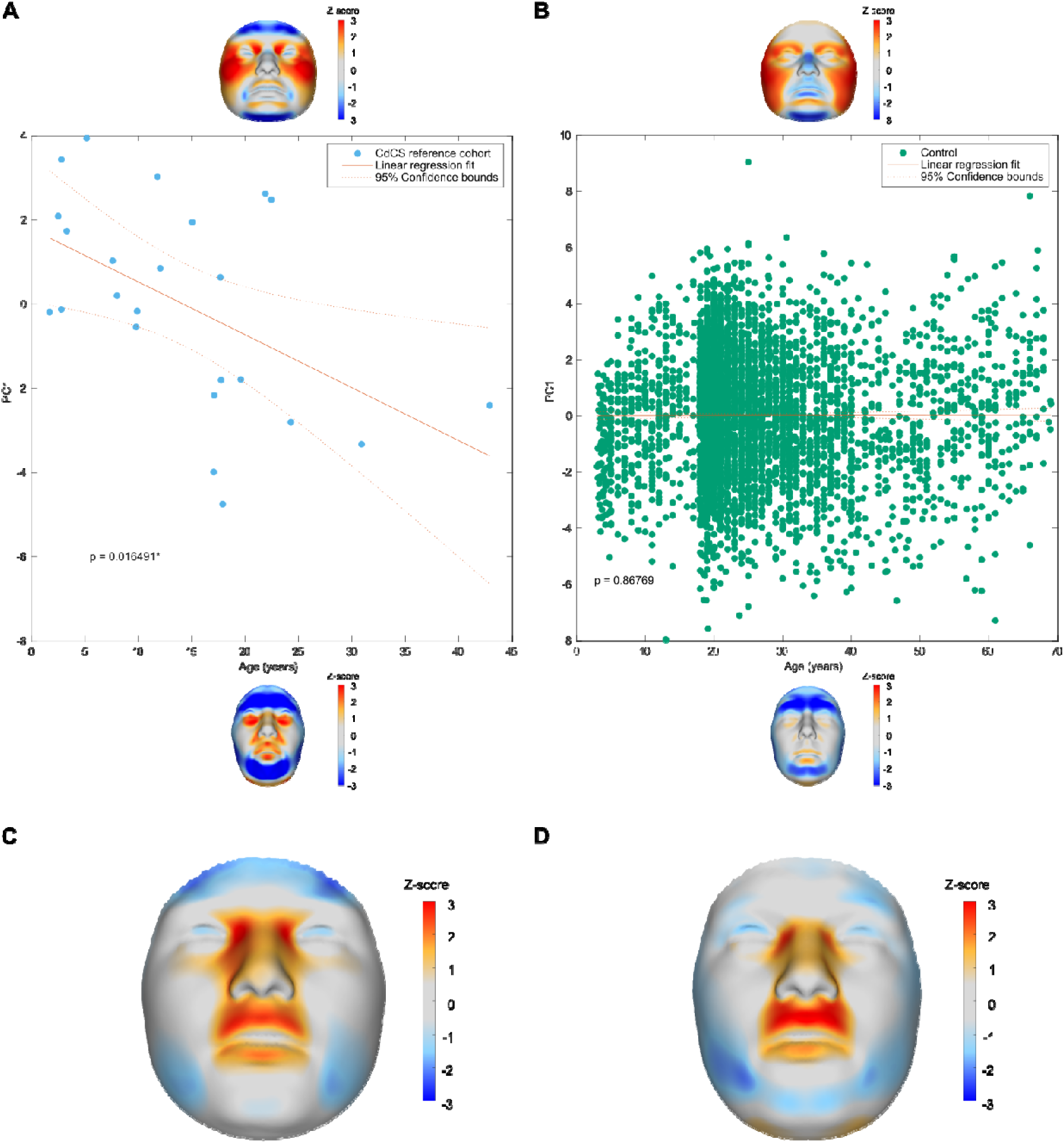
Age effect. **(a)** Linear regression of age on the first principal component from a PCA on age-adjusted shapes of the Cri-du-Chat Syndrome (CdCS) cohort **(b)** and age-adjusted shapes of the control sample. The facial shapes and signatures at +- 3 standard deviations of the PCs are shown. **(c)** Mean facial signature of the twelve youngest participants of the CdCS cohort and **(d)** the twelve oldest participants.

**Supplementary Figure 8:**
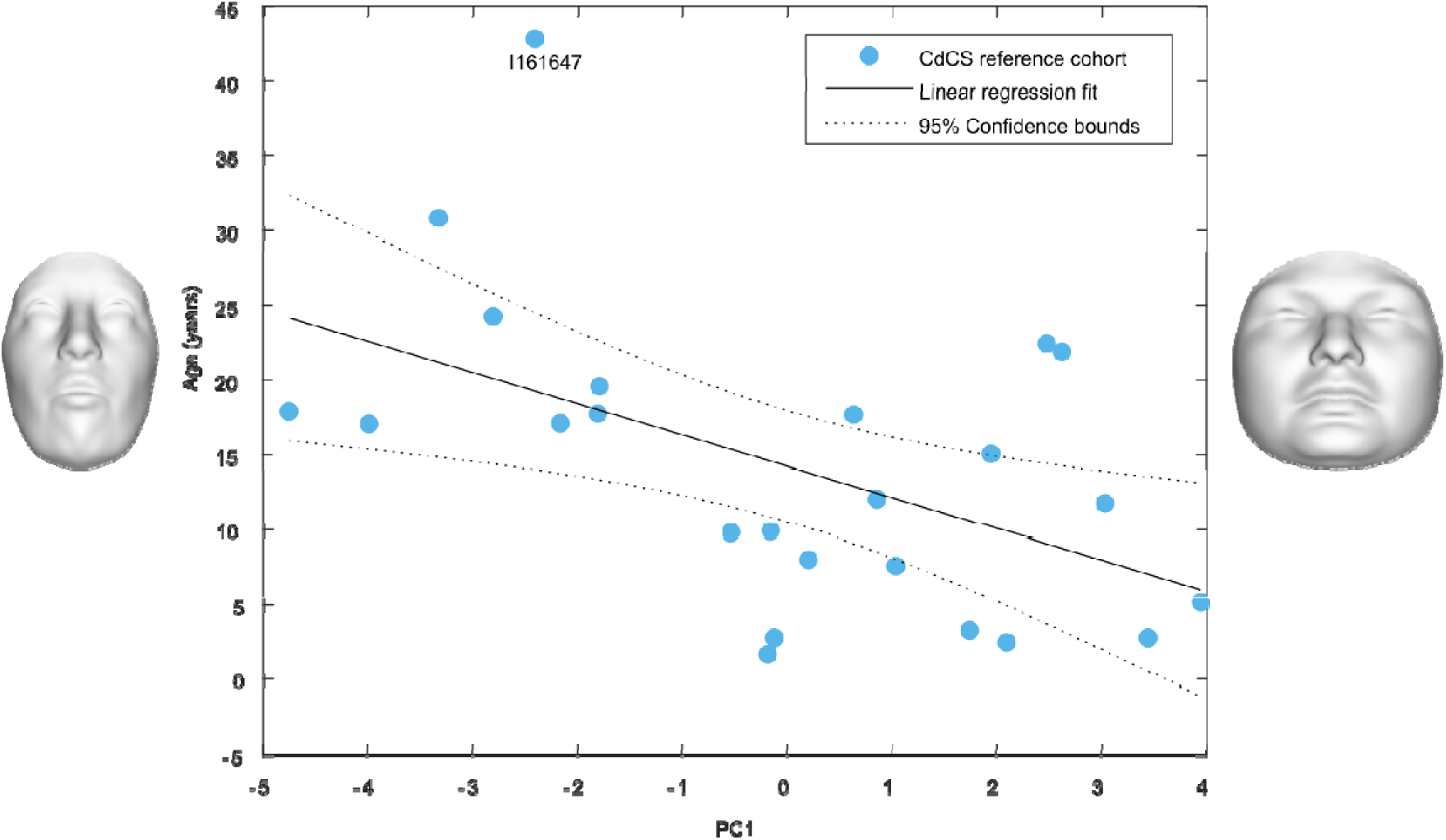
Age outlier. Linear regression of the first principal component on age from a PCA on age-adjusted shapes of the Cri-du-Chat Syndrome (CdCS) cohort, with participant I161647 annotated. The facial shapes at +- 3 standard deviations of PC1 are shown.

