## Supplementary Media 1-3 for "Advancing Genotype-Phenotype Analysis through 3D Facial Morphometry: Insights from Cri-du-Chat Syndrome"

### Slide 1
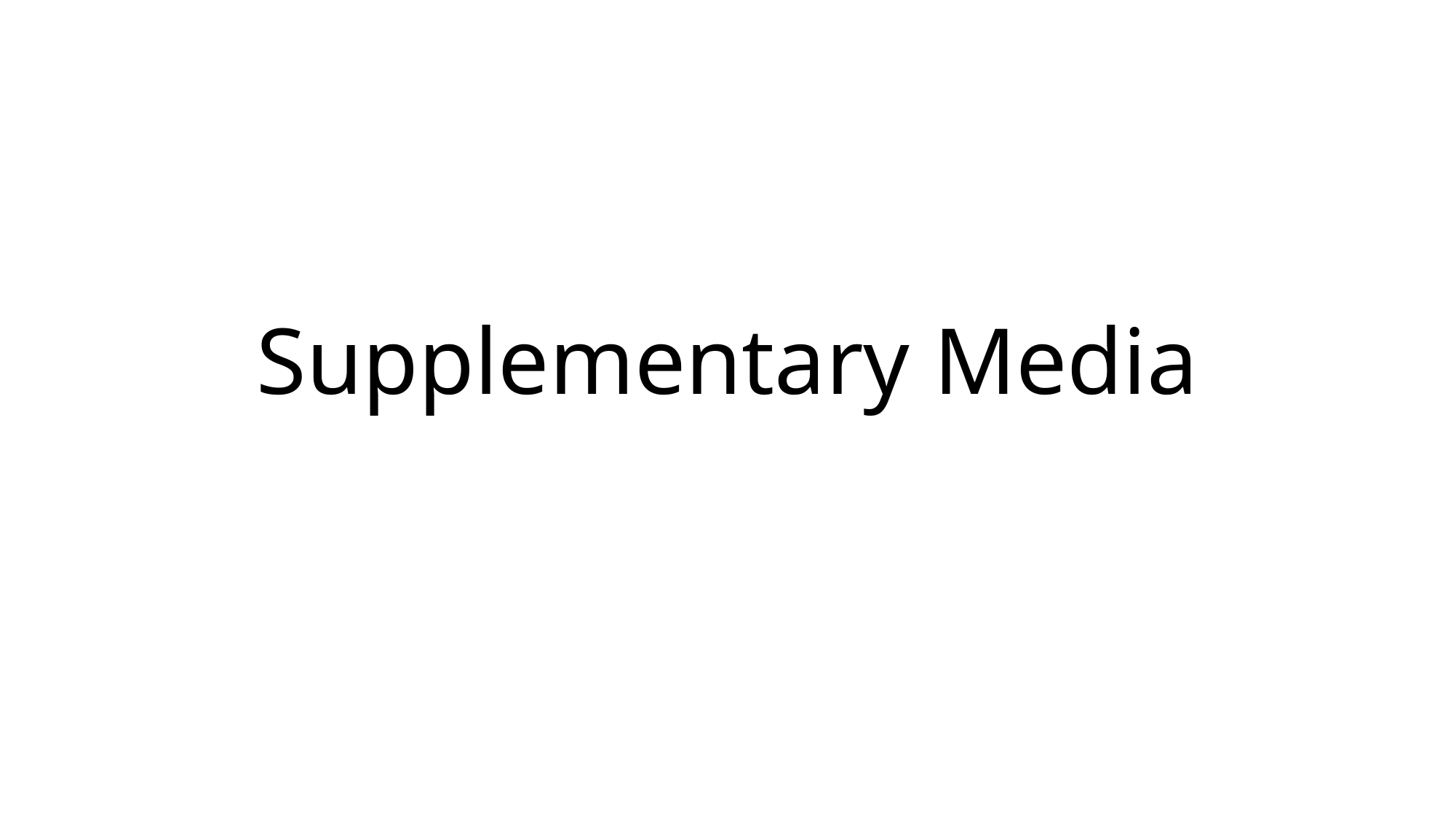

#### Supplementary Media

### Slide 2
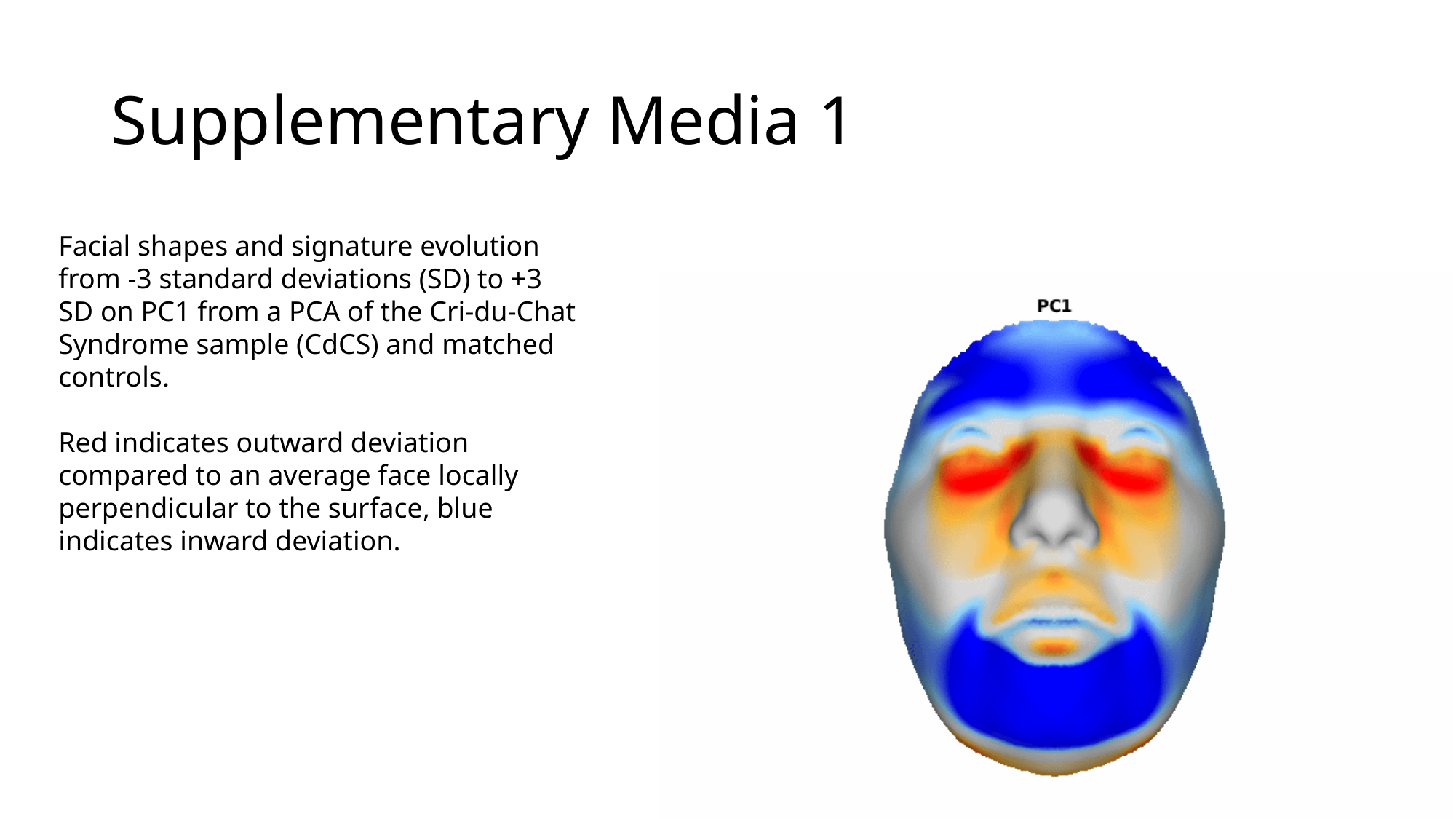

#### Supplementary Media 1
Facial shapes and signature evolution from -3 standard deviations (SD) to +3 SD on PC1 from a PCA of the Cri-du-Chat Syndrome sample (CdCS) and matched controls.
Red indicates outward deviation compared to an average face locally perpendicular to the surface, blue indicates inward deviation.

### Slide 3
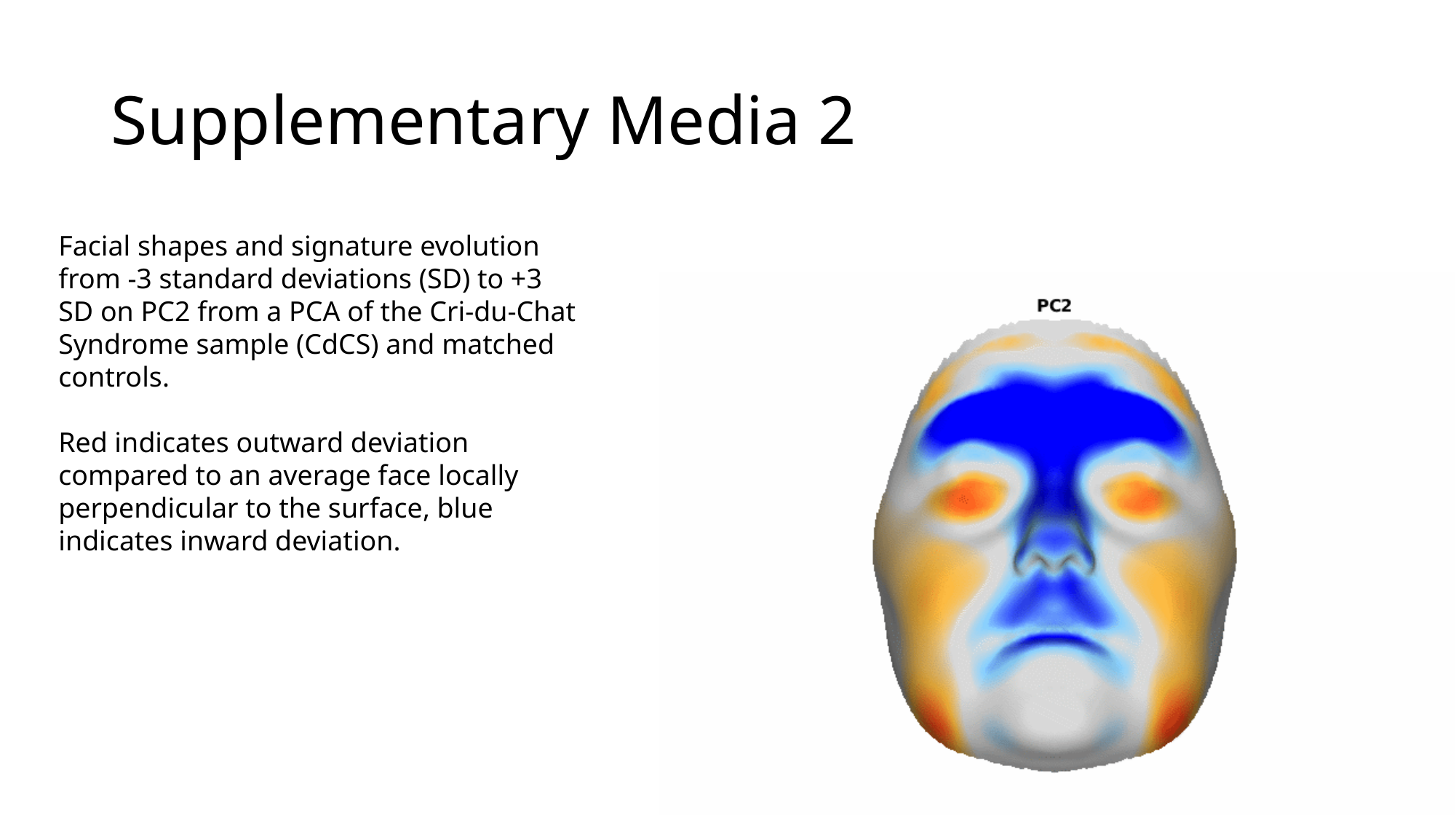

#### Supplementary Media 2
Facial shapes and signature evolution from -3 standard deviations (SD) to +3 SD on PC2 from a PCA of the Cri-du-Chat Syndrome sample (CdCS) and matched controls.
Red indicates outward deviation compared to an average face locally perpendicular to the surface, blue indicates inward deviation.

### Slide 4
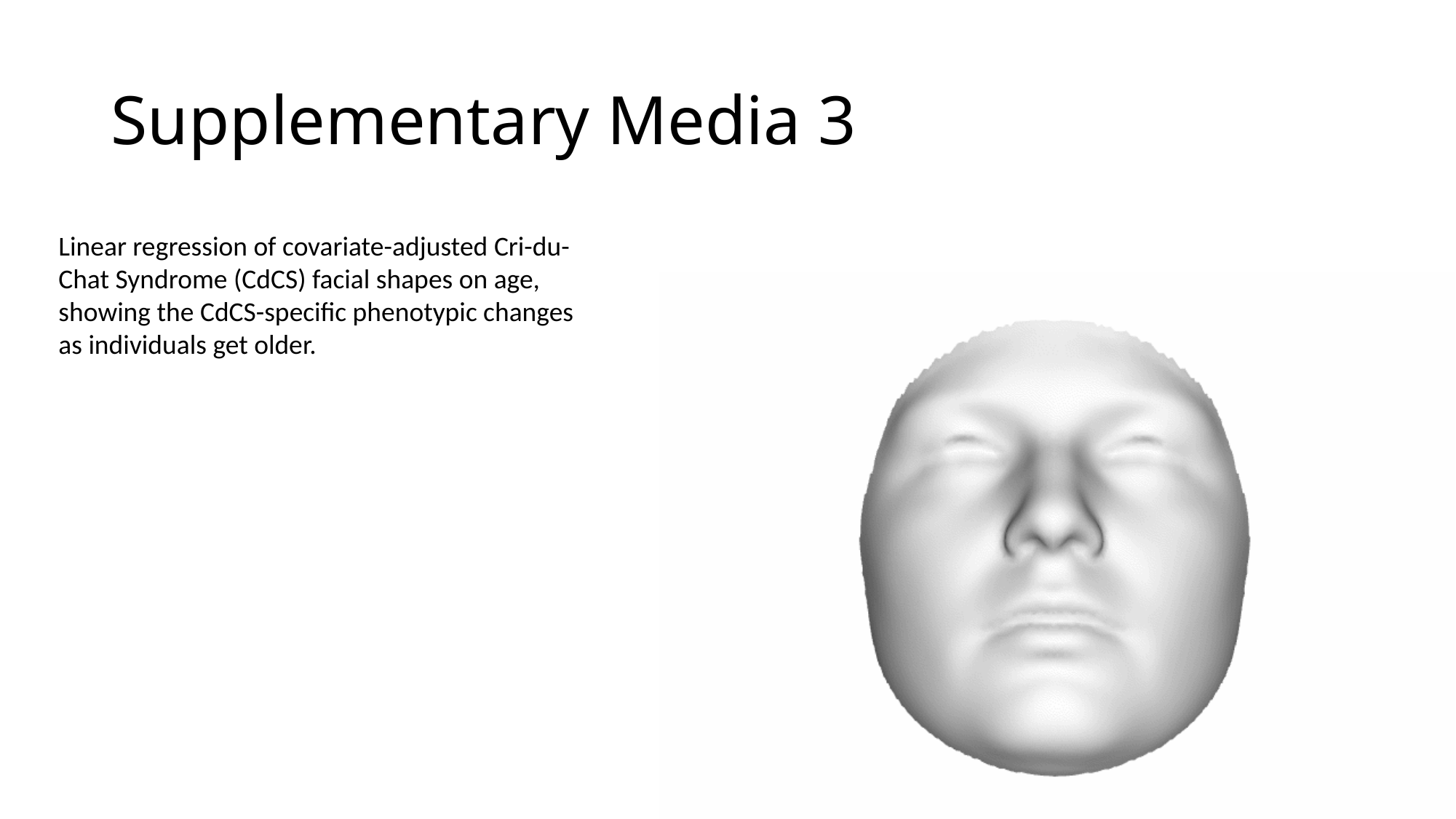

#### Supplementary Media 3
Linear regression of covariate-adjusted Cri-du-Chat Syndrome (CdCS) facial shapes on age, showing the CdCS-specific phenotypic changes as individuals get older.
